# Hypovitaminosis D: How Common is it, and what are its Predictors among Children with Congenital Heart Disease Attending the Lagos State University Teaching Hospital?

**DOI:** 10.64898/2025.12.23.25342696

**Authors:** Lawani O Faith, Animasahun B Adeola, Adekunle O Motunrayo, Goodness A Animasahun, Ariyibi A Adedayo, Hughes-Darden Cleo

## Abstract

Malnutrition is a deficiency of both macro- and micro-nutrients. Malnutrition is common in children with congenital heart defects due to reduced intake, poor gastrointestinal absorption resulting from gut hypo-perfusion and increased metabolism. The present study aimed to determine the prevalence and predictors of hypovitaminosis D in children with congenital heart defects aged 1 to 12 years compared with apparently healthy controls.

**Methods:** A comparative cross-sectional study conducted from July to November 2020 involving 115 children with congenital heart disease and 115 apparently healthy controls matched for age, sex and socio-economic class. A self-designed proforma was used to collect information on subjects’ bio-data, socio-economic class, health information, morbidity pattern, frequency and duration of sunlight exposure and 48 hours’ dietary recall of vitamin D containing foods. Weight, height, waist and hip circumference were measured, BMI was calculated, waist hip ratio, weight-for-age, height-for-age and BMI-for-age Z scores were obtained. Blood samples were collected for measurement of serum 25-hydroxyvitamin D levels. Hypovitaminosis D was taken as both vitamin D deficiency and insufficiency with serum 25(OH)D levels of less than 20ng/ml and 20.1-29.9ng/ml respectively. Descriptive and inferential analysis was carried out using SPSS version 27.

**Result:** The prevalence of hypovitaminosis D in subjects was 59.1% with median (interquartile range) of 28.13ng/ml (21.2-42.3ng/ml) while the prevalence in healthy controls was 41.7%. The difference was statistically significant (p= 0.008). There was no statistically significant difference in the prevalence of hypovitaminosis D between the acyanotic and cyanotic subjects. The age groups greater than 3 years, female sex and duration of diagnosis greater than 48 months were independent predictors of hypovitaminosis D in subjects.

**Conclusion:** children with congenital heart defect should have their serum 25-hydroxyvitamin D measured at intervals and may benefit from treatment if found to have vitamin D deficiency.

## Introduction

Congenital heart defects (CHD) are the commonest birth defects worldwide, affecting millions of newborn babies every year and accounting for nearly one-third of all major congenital anomalies.^1^ They are typically defined as structural abnormalities of the heart and the great vessels present at birth.^2^ Congenital heart defects contribute to childhood morbidity and mortality especially in developing countries where intervention facilities are readily not accessible and available.^3^

The estimated worldwide prevalence of congenital heart defects is generally considered to be 8 per 1000 live births.^4^ However, this estimate is perhaps inaccurate and does not take into consideration regional differences and the increasing access to healthcare and diagnostic technologies.^2^ Otaigbe and Tabansi^5^ reported a prevalence of 14.4 per 1000 children in the Niger delta region of Nigeria from a hospital based study while Ujuanbi et al^6^ reported a prevalence of 18.1 per 1000 children from a population based study also from the Niger delta region of Nigeria. In both studies, the high prevalence was attributed to the increasing oil spillage and gas flaring from petroleum exploitation in this region. Previous reports on the incidence and prevalence of congenital heart defect in Nigeria ranged between 3.5 and 4.6 per 1000 births respectively.^7,8^ A recent prevalence study carried out among neonates by Ige et al^9^ though a hospital based study in north-central Nigeria reported a prevalence as high as 28.8 per 1000 live births.

Congenital heart defects are a major cause of mortality in children with congenital malformations and most congenital heart defect deaths (greater than 90%) occur within the first five years of life, mainly during infancy.^10–13^ Children with congenital heart defects are prone to several complications including malnutrition.^14–16^ Malnutrition (deficiency of both macro- and micro-nutrients) occurs as result of inadequate nutrition caused by or due to feeding difficulties, poor nutritional absorption from the digestive tract in those with chronic congestive heart failure, increased caloric requirement to sustain the increased myocardial, respiratory and neuro-humoral functions in congenital heart defect related heart failure.^14^ Malnutrition in children with congenital heart defect is a major contributor to their morbidity and mortality.^15^ Malnutrition increases the risk of frequent hospitalization, contributes to the development of heart failure, a common presentation in affected children. It also worsens the morbidity and mortality following corrective surgical operations and postoperative recovery. Hassan et al^16^ in a University Children’s Hospital in Egypt and Okoromah et al^14^ in a tertiary hospital in Lagos, Nigeria reported a high prevalence of macronutrient malnutrition of 84% and 90.4% respectively in children with congenital heart defects.

Vitamin D is a known micronutrient and the effects of its deficiency has been of global interest in recent times attracting the attention of researchers and clinicians. Children with congenital heart defects are at a greater risk of hypovitaminosis D due to the mechanisms earlier mentioned and to the reduced exposure to sunlight.^17^ Children with cyanotic congenital heart defects are more likely to be malnourished as compared with those with acyanotic congenital heart defect due to their reduced oxygen saturation as chronic hypoxia impairs cellular metabolism and cell growth.^16,18,19^ Few studies done outside Africa have reported a high prevalence of vitamin D deficiency (55.3% - 70.3%) in children with CHD.^17,20^ Vitamin D has been recognized as an important vitamin for cardiovascular health and its deficiency as a potential risk factor for several cardiovascular disease processes.^21–24^ Vitamin D is also important in immune system function through its anti-inflammatory properties hence its deficiency is associated with increased risk of infections such as upper respiratory tract infections.^25^

There is no report of the prevalence of hypovitaminosis D in children with congenital heart defects in Nigeria and the West African sub-region despite the high burden of congenital heart defect in the region known to the present researcher. The burden of hypovitaminosis D in children with congenital heart defects in Nigeria may differ from those published from other regions of the world due to differences in the availability of sunlight and fortification of food which is widely adopted in developed countries.^26^ Findings from the proposed study will help to identify the burden of this nutrient deficiency and provide the opportunity for correction of this deficiency in the study population. It will guide on establishing protocols for routine screening and prompt treatment which will thereby contribute to reduction in morbidity and mortality of children with congenital heart defects.

## Methods

This was an analytical cross-sectional conducted over five months (1^st^ of July 2020 to 30^th^ of November 2020) at the Lagos State University Teaching Hospital (LASUTH), Nigeria. LASUTH is a state-owned tertiary center that accommodates referrals from all parts of Lagos state and its neighbouring states.^27,28^ Its Paediatric unit has 80 beds, wards and emergency rooms inclusive.^27,28^ The paediatric cardiology clinic attends to all cardiology cases from different ethnic groups, socioeconomic class and across religious divides.

One hundred and fifteen children aged one to twelve years who had previously been diagnosed with congenital heart disease after an echocardiography and were attending the paediatric cardiology clinic. The study population also included apparently healthy controls from the paediatric out-patient clinic who were matched for age, sex and socio-economic class whose parents/guardian gave a written informed consent. Exclusion criteria in the study include Children with overt signs of rickets, Children with features of liver disease. Children with features suggestive of sickle cell anaemia. Children with features of renal disease e.g. oliguria, facial swelling, frothy urine, Children on vitamin D or multivitamin supplements in the last 2 months (half-life of 25(OH)D is 3 weeks), Children on anti-convulsant drugs and steroids. Ethical approval was obtained from Health Research Ethics Committee of Lagos State University Teaching Hospital

Written and verbal informed consent were obtained from all parents/caregivers while an assent form was filled by children who were 7 years of age and above. Data was collected and entered by the researcher. The study participants were assigned with numeric codes for identification.

The cost of the investigation was borne by the investigator.

On recruitment, general clinical examination, anthropometry and social class classification were carried out for each consenting participants Information on duration and frequency of sunlight exposure, type and frequency of intake of vitamin D containing foods in the last 48 hours before recruitment were also assessed among participants.

Classification of subjects’ nutritional status was based on WHO criteria. The WHO recommends the use of standard definitions and classifications for malnutrition based on calculated Z scores for anthropometric indices.^29^ BAZ greater than or equal to −2 to +1 was defined as normal weight, BAZ greater than +1 to less than or equal to +2 as overweight, BAZ greater than +2 to +3 as obese, BAZ greater than +3 as severely obese, BAZ less than −2 as wasted and BAZ less than −3 as severely wasted. WAZ less than −2 was defined as underweight and less than −3 as severely underweight and HAZ less than −2 as stunted and less than −3 as severely stunted.^29,30^

Two milliliters of venous blood was collected from subjects and the blood collected was placed in a plain bottle and allowed to clot as serum is needed for the assay. At the research laboratory, samples were centrifuged the plasma stored at −80’C.^31,32^ Frozen samples were allowed to thaw and equilibrate to room temperature for 30 minutes before use. Samples were maintained at room temperature by not keeping samples under direct sunlight or a heat source. Samples were analyzed in the chemical pathology laboratory by a chemical pathologist and the researcher using a 25-hydroxyvitamin D ELISA kit by Calbiotech®^31^, manufactured in California, United States of America.

### Data Analysis

Analysis of data was done using the SPSS version 23.0 (IBM version 26, Illinois, United State of America).

Categorical data were presented as frequencies and percentages while numeric data were represented using mean and standard deviation, median and interquartile range were used to describe skewed data. Kolmogorov Smirnov test was used to assess data normality distribution. Chi square or Fischer’s exact test were used to test the association between categorical variables. Independent Student t test or Mann Whitney U test if skewed were used to compare numeric data. Logistics regression analysis was used to determine independent factors associated with vitamin D deficiency. P-value <0.05 was taken as statistically significant at 95% Confidence Interval (CI).

## Results

A total of 230 children aged one to twelve years were recruited for the study. Out of this number, 115 were children with congenital heart disease and 115 were apparently healthy children (controls) who were matched for age and sex and socio-economic class.

Table I shows the socio-demographic and anthropometric distribution of all participants. There was no statistically significant difference between the subjects age, gender, social class, ethnic group and religion and those of healthy controls. The mean age for cases was 4.77 ± 1.4 years and the mean age for controls was 4.74 ± 1.4 years. The male-to-female ratio of participants was 1.2:1. Average HAZ, WAZ BAZ and Waist-hip ratio were higher in controls than the subjects (*p* < 0.001).

**Table I:** Socio-demographic and anthropometric characteristics of participants.

| | Subject(n=115)<br>n(%) | Control(n=115)<br>n(%) | Total | $\chi^2$ | p-value |
| --- | --- | --- | --- | --- | --- |
| Age group (Years) |  |  |  |  |  |
| 1-3 | 43 (37.4) | 43 (37.4) | 86 (37.4) | 0.000 | 1.000 |
| 4-6 | 37 (32.2) | 37 (32.2) | 74 (32.2) |  |  |
| 7-9 | 21 (18.2) | 21 (18.2) | 42 (18.2) |  |  |
| 10-12 | 14 (12.2) | 14 (12.2) | 28 (12.2) |  |  |
| Mean ± SD | 4.77 ± 1.4 | 4.74 ± 1.4 | | 0.019* | 0.934 |
| Gender |  |  |  |  |  |
| Male | 64 (55.7) | 64(55.7) | 128 (55.7) | 0.000 | 1.000 |
| Female | 51 (44.3) | 51(44.3) | 102 (44.3) |  |  |
| Social class |  |  |  |  |  |
| Lower | 25 (21.7) | 25 (21.7) | 50 (21.7) | 0.000 | 1.000 |
| Middle | 35 (30.4) | 35 (30.4) | 70 (30.4) |  |  |
| Upper | 55 (47.8) | 55 (47.8) | 110 (47.8) |  |  |
| HAZ | -2.20 ± 1.9 | 1.24 ± 2.1 | | 3.708 | <0.001* |
| WAZ | -1.21 ± 1.8 | 0.14 ± 1.4 | | 6.221 | <0.001* |
| BAZ | 0.29 ± 1.9 | 1.35 ± 1.1 | | 5.152 | <0.001* |
| Waist hip ratio | 0.92 ± 0.1 | 1.01 ± 0.1 | | 8.528 | <0.001* |

Table II shows the echocardiographic diagnosis of subjects. The acyanotic and cyanotic CHD comprised 61.7% and 38.3% respectively of the CHD subjects with a ratio of 1.6:1. Ventricular septal defect was the commonest acyanotic CHD while tetralogy of Fallot accounted for the most common cyanotic heart defect. Twenty-seven (23.5%) of the subjects had been previously hospitalized. Out of this, 81.5% of them were admitted once. Up to 35 (30.4%) of subjects were on medications. The majority of those on medication (52.8%) were on oral propranolol. The subjects had been diagnosed with congenital heart disease for a median (interquartile range) duration of 25 (14 to 47) months

**Table II:**
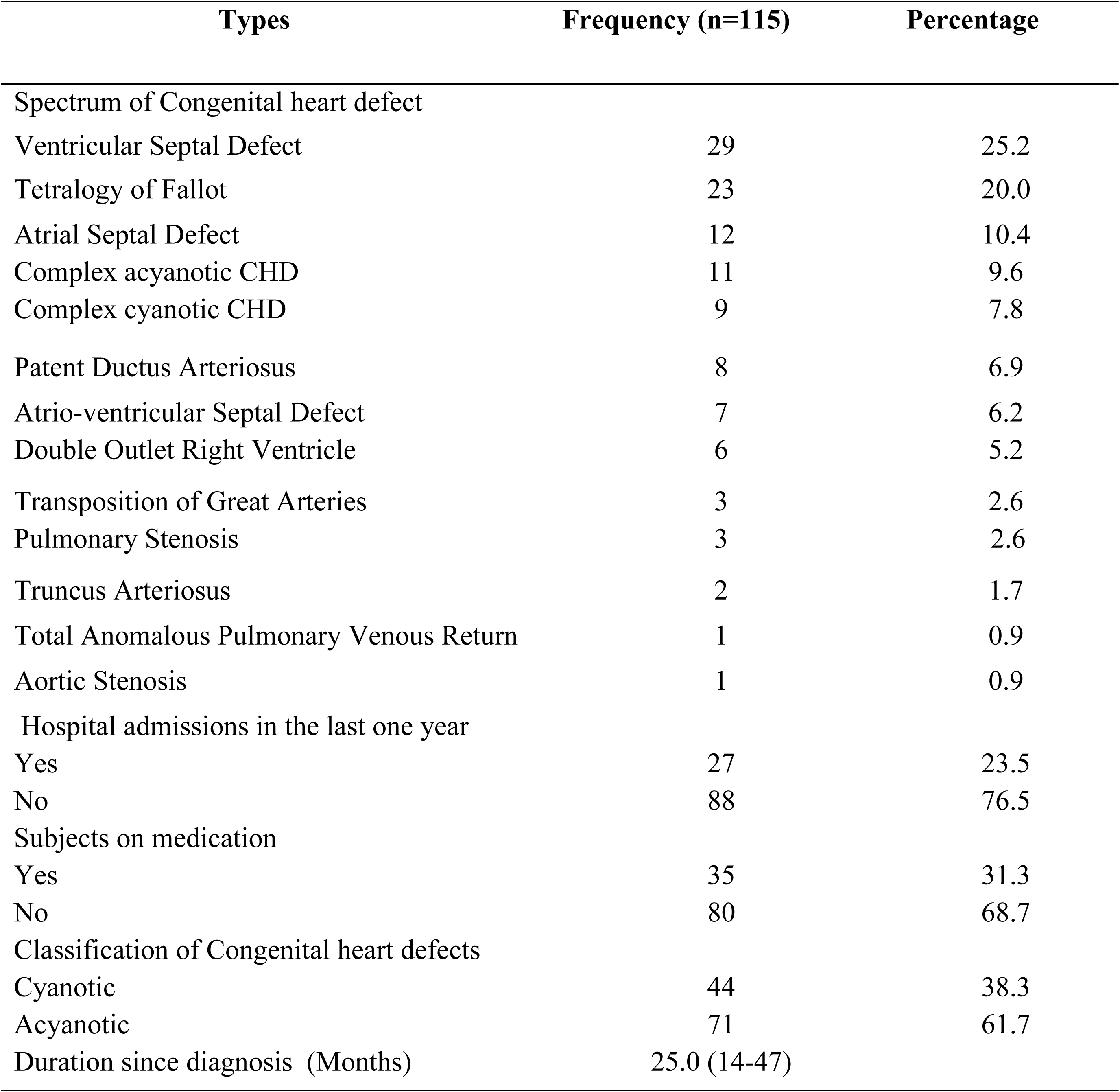
Spectrum of Congenital heart defects and clinical characteristics among subjects.

The difference in the prevalence of hypovitaminosis D in the subjects was statistically significantly higher (59.1%) than 41.7% in the controls (*p=* 0.008). Figure 1 shows that there were three times more children with vitamin D deficiency among subjects than there were in the controls (24.3% vs 7.8%,□^2^ =13.28, *p=* <0.001). children with CHD had significantly lower median (inter quartile range) serum levels of serum 25-hydroxyvitamin D levels compared to controls [28.13(21.2-42.3) versus 31.62(26.0-52.9) *p*= 0.004].

**Figure 1:**
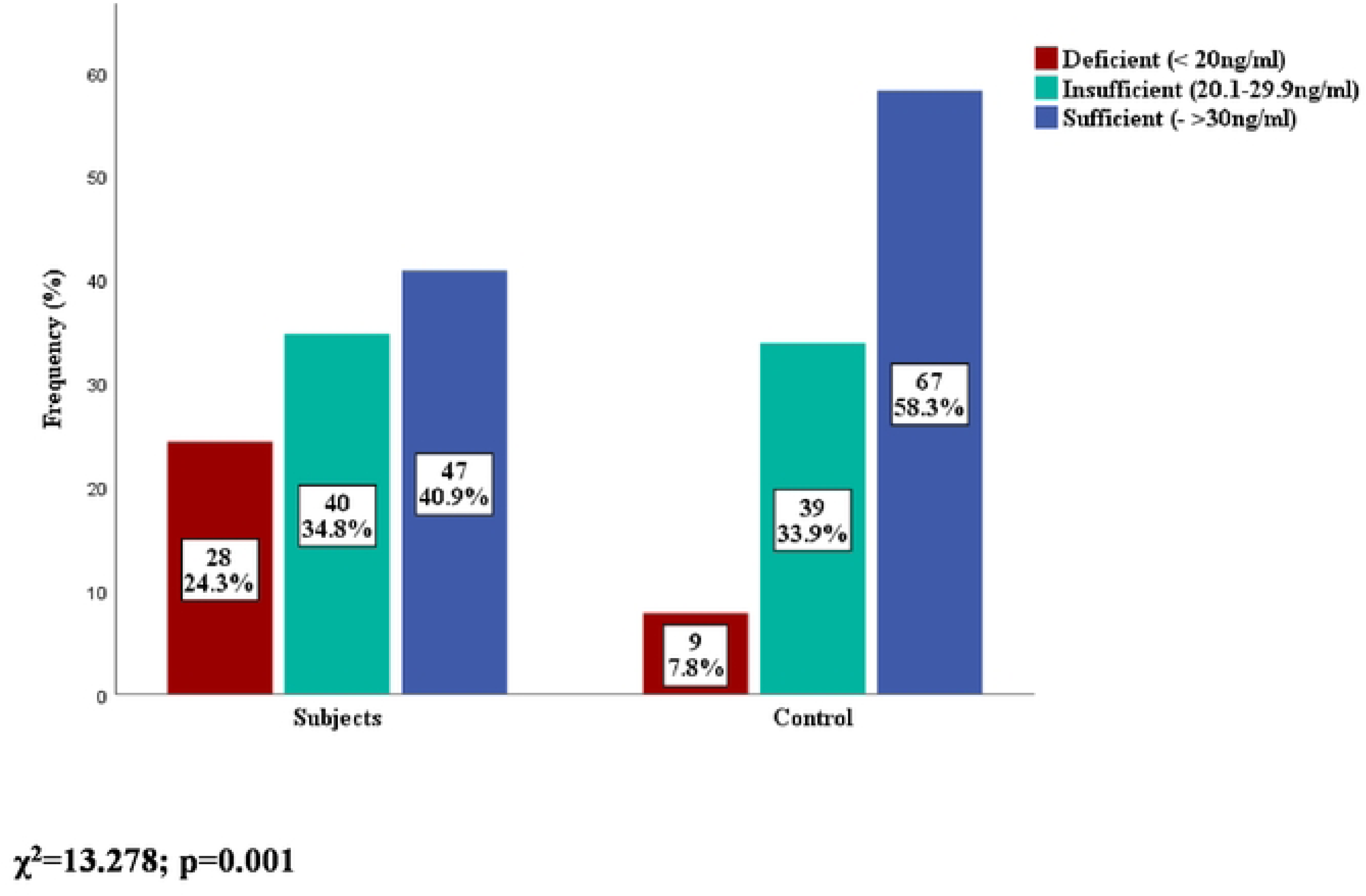
Serum vitamin D status among subjects and controls.

Table III shows the relationship between the selected factors and vitamin D status among subjects. Hypovitaminosis D was statistically significantly more prevalent in older age groups compared with the age group of 1-3 years (*p* < 0.001). The prevalence of hypovitaminosis D was also significantly higher in females as compared to males (*p* = 0.009). There was however no significant association between socio-economic class and hypovitaminosis D. The subjects whose diagnosis was made more than 24 months before recruitment had a significantly higher prevalence of hypovitaminosis D compared with those whose were less than 24 months (*p* < 0.001). The prevalence of hypovitaminosis D was significantly much higher in those whose duration since diagnosis was above 48 months. A higher proportion of subjects not on medication had statistically significant hypovitaminosis D compared with those on medication (*p =* 0.031). Previous hospitalization and frequency of hospitalization had no statistically significant relationship with hypovitaminosis D. The prevalence of hypovitaminosis D among subjects with stunting, underweight and abnormal categories of body mass index was not statistically significant between vitamin D deficient and vitamin D sufficient subjects.

**Table III:**
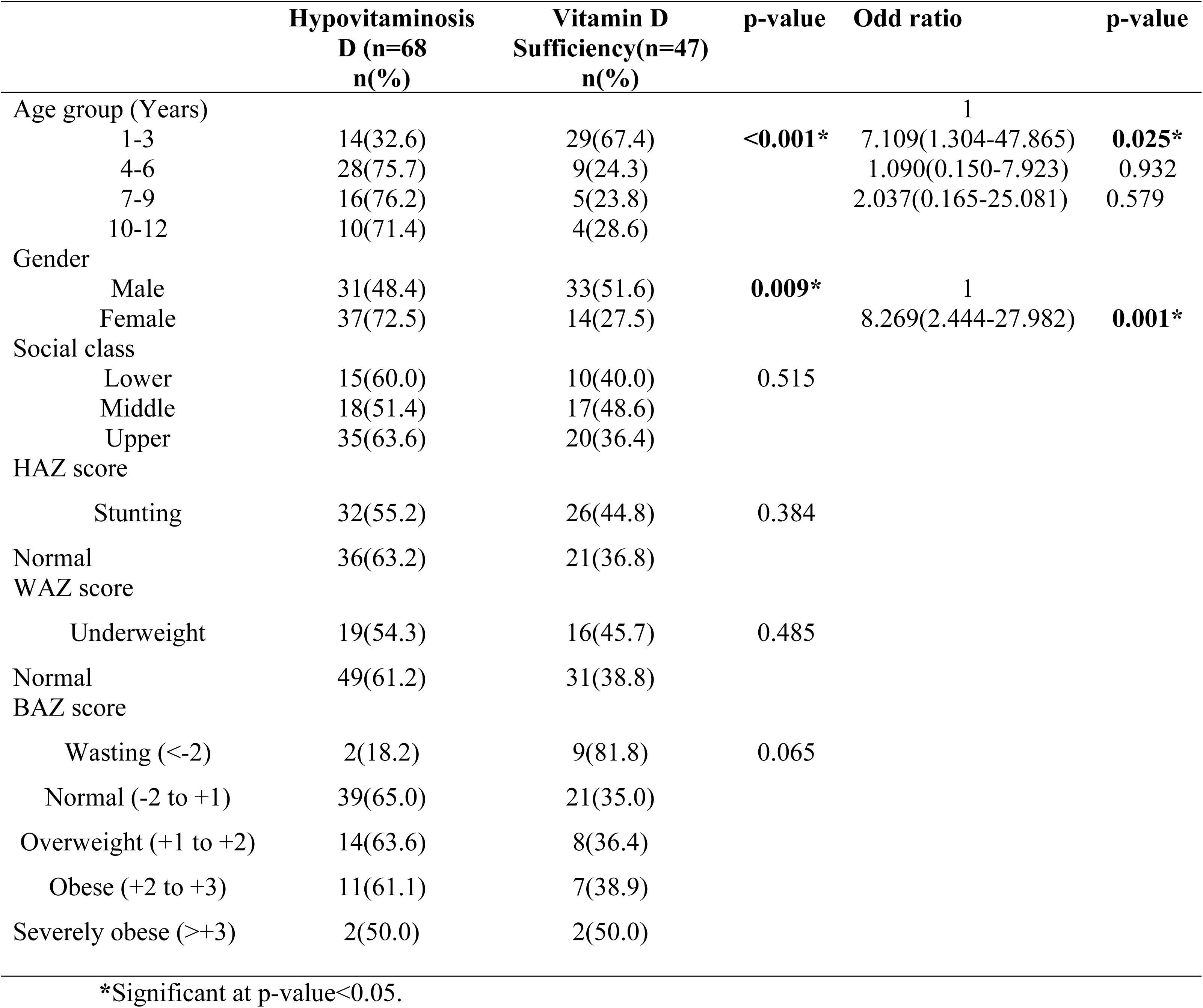
The relationship between vitamin D status and socio-demographic/anthropometric characteristics of subjects.

**Table IV:**
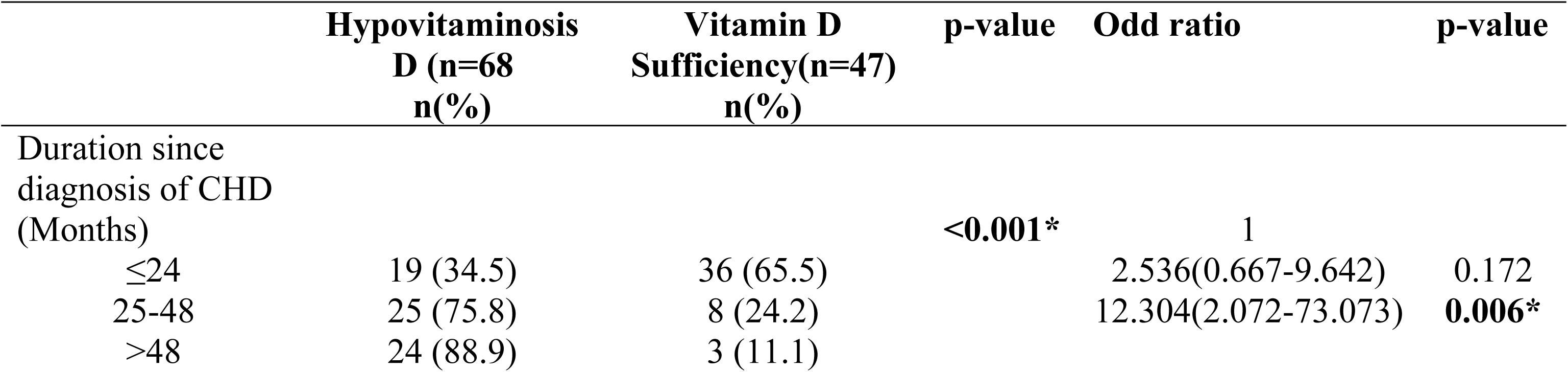
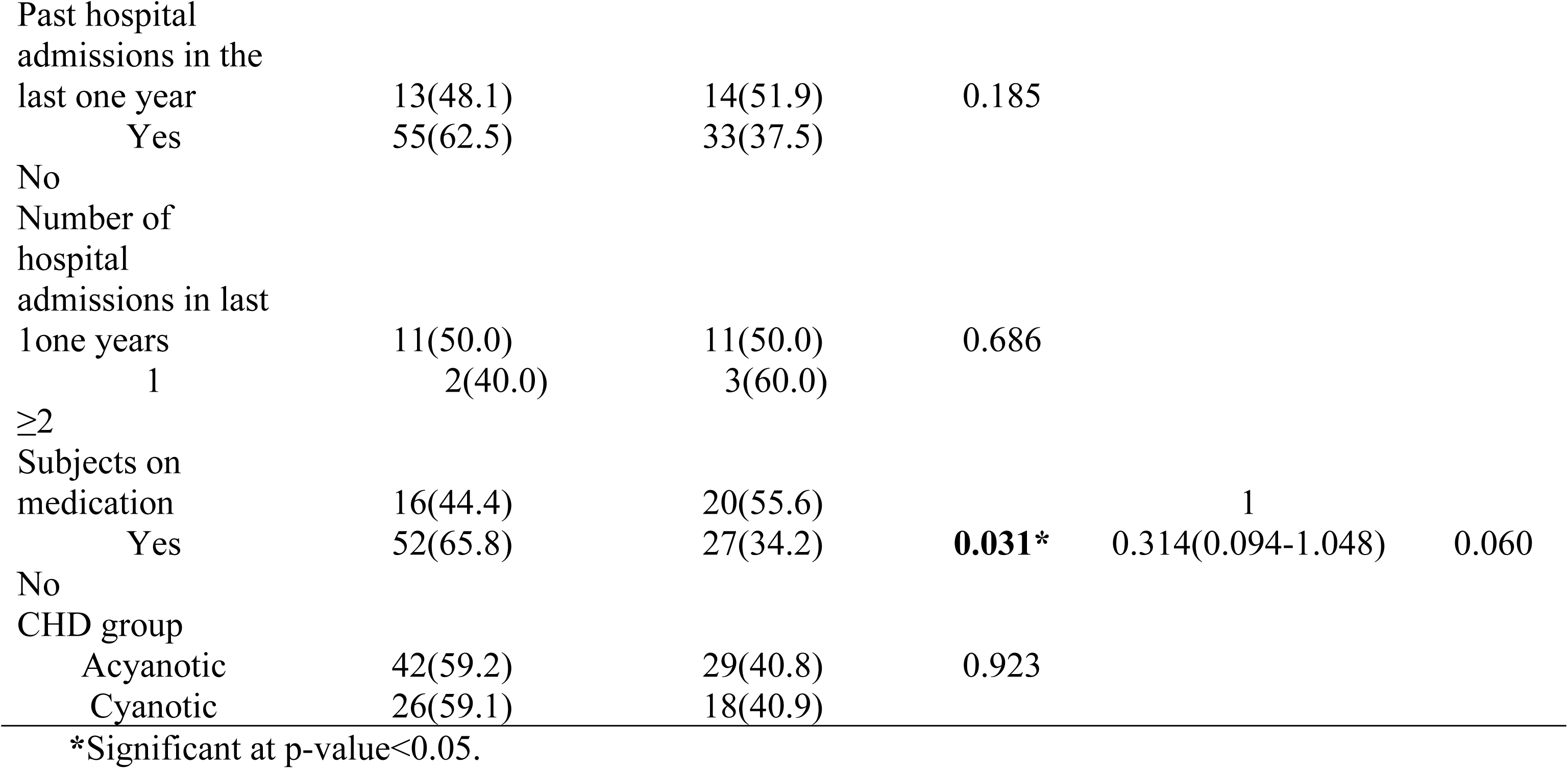
The relationship between vitamin D status and clinical characteristics of subjects.

Multivariable logistic regression to determine the independent predictors of hypovitaminosis D in subjects. Analysis showed age between 4-6 years, female sex and subjects who had been diagnosed with congenital heart disease for greater than 48 months before recruitment were independent predictors of hypovitaminosis D.

## Discussion

The current study aimed to determine the prevalence and the predictors for hypovitaminosis D in children with congenital heart defect attending the Lagos State University Teaching Hospital. The prevalence of vitamin D deficiency recorded in the present study was 24.3%. This is lower than in the studies by Passeri^17^ in Italy and McNally^33^ in Canada who found 55.3% and 46% respectively of children with congenital heart disease to be vitamin D deficient but the proportion of those who had vitamin D insufficiency were not specified in the latter studies. The above difference in the prevalence of vitamin D deficiency despite similar cut off values for hypovitaminosis D could be due to the fact that children less than one year were excluded from the present study and included in the studies by Passeri^17^ and McNally.^33^ Children less than one year of age have been shown to have falsely elevated levels of serum 25-hydroxyvitamin D due to the presence of C3-epi25-hydroxyvitamin D3, an epimer of vitamin D3 due to hepatic immaturity.^34^ So the inclusion of infants may have falsely elevated the proportion of subjects with vitamin D deficiency in the studies by Passeri and McNally. This might also explain the nearly four-fold levels of vitamin D deficiency found in neonates in the study by Graham^35^ in the United States of America when compared with the present study. A better comparison of this prevalence would have been with studies done in Africa and thus Nigeria that excluded children less than one year of age.

The serum levels of 25-hydroxyvitamin D in children with congenital heart disease was compared with that of apparently healthy controls in the present study. Children with congenital heart disease were seen to have significantly higher levels of vitamin D deficiency. Vitamin D deficiency is defined by the US Endocrine Society as serum 25-hydroxyvitamin D levels less than 20ng/ml. Noori^36^ and Salehi^37^ in Iran also recorded a similar finding where children with congenital heart disease had significantly higher levels of vitamin D deficiency compared with healthy controls.

There was no significant difference in the prevalence of hypovitaminosis D between children with acyanotic and cyanotic congenital heart disease. This finding is similar to the study by Salehi et al^37^ among Iranian children with congenital heart disease but contrasts with the study by Noori et al^36^ where the children with cyanotic congenital heart disease had significantly lower levels of serum 25-hydroxyvitamin D. Chronic hypoxia is seen in children with cyanotic congenital heart disease and the effect of this hypoxia makes them more prone to malabsorption due to poor intestinal perfusion.^16,18,19^ A plausible reason why this did not reflect in the present study may be because the children with cyanotic congenital heart disease were in the younger age group and thus the hypoxia associated with this condition may not yet have become chronic.

Hypovitaminosis D was seen to be significantly commoner in children between four and twelve years than in younger participants in this study. This is similar to findings by Passeri^17^ and Noori^36^ where older children had more vitamin D deficiency. Even in apparently healthy children, this trend has been observed.^38^ This is because with increasing age, the amount of 7-dehydrocholesterol in the skin which is needed for the formation of cholecalciferol by sunlight decreases and this reduces the synthesis of vitamin D.^39,40^ Another plausible reason maybe that younger children consume more milk and eggs and have more sunlight exposure and this could be related to socio-cultural factors in our environment where the meals of most younger children are milk based and their mothers run errands with them and thus are more exposed to sunlight. With increasing industrialization and modernization in African communities, older children are likely to spend more time indoors having increased screen time and other activities that favour a sedentary lifestyle.^41^

Hypovitaminosis D was significantly commoner in females than in males. This finding is dissimilar to studies by Passeri^17^, Noori^36^ and Salehi^37^ where there was no significant difference in the serum levels of 25-hydroxyvitamin D in both sexes. Passeri^17^ however reported more females had severe forms of vitamin D deficiency although their finding was not significant. Females could be at a greater risk for hypovitaminosis D compared with their male counterparts as females tend to engage less in outdoor activities and hence have lesser sunlight exposure compared with males. Another plausible reason for lower levels of vitamin D in females may be due to the lower levels of the female hormone, oestrogen in pre-adolescents. Oestrogen and vitamin D mutually affect each other’s biosynthesis. Oestrogen stimulates the synthesis of 1,25-(OH)D in the kidneys as both of them regulate each other’s receptor expression.^42^ Therefore, lower levels of oestrogen will result in reduced production of vitamin D.

There was no significant difference in the levels of serum 25-hydroxyvitamin D among the different socioeconomic classes. Family income directly affects nutritional status and thus children whose parents are in the low socioeconomic class may have less access to vitamin D rich foods.^43^ In the general population, children whose parents are in the low socioeconomic class have been shown to have higher levels of serum 25-hydroxyvitamin D than those in the middle and upper socioeconomic classes due to lesser sunlight exposure by children in the latter socioeconomic classes.^44,45^ Children with congenital heart defect, irrespective of their socio-economic class have lesser sun exposure due to the chronic nature of their illness and this may possibly explain why socio-economic class was not associated with hypovitaminosis D. Consumption of milk with meals and locally prepared cereals was associated with significantly higher levels of 25-hydroxyvitamin D in the present study. These are food items that are easily available and affordable and this might be a reason why there was no significant difference among the socioeconomic groups. In the last decade or so, milk manufacturers in Nigeria have been packaging milk in small affordable sachets making it more available.

Less than 10% of vitamin D is derived from food sources. Production of vitamin D is majorly dependent on sunlight exposure. Thus, it would be expected that subjects who had more exposure to sunlight (frequency and duration) would have had higher levels of serum 25-hydroxyvitamin D, but this was not reflected in the present study. Exposure to sunlight does not necessarily translate to increase in serum 25-hydroxyvitamin D in blacks due to the presence of melanin in the skin which when converted to eumelanin inhibits the penetration of ultraviolet B in the skin thus reducing vitamin D production.^46^Hence, levels of serum 25-hydroxyvitamin D might not be majorly dependent on sunlight exposure in the tropics and thus further supports the need for food fortification practices.

Subjects whose diagnosis of congenital heart disease was made more than 48 months before recruitment had significantly higher prevalence of hypovitaminosis D. In India, Vaidyanathan^47^ reported that older age at corrective intervention in children with congenital heart disease was a significant predictor for malnutrition at presentation and early surgical correction of the cardiac anomaly favourably influences the nutritional status of children with congenital heart disease. This might be because the longer these defects persist, the longer the haemodynamic abnormalities which can result in gut hypo-perfusion and hence predispose them to malnutrition including micro-nutrient deficiency like hypovitaminosis D which was reflected in the present study.

The present study showed that higher proportion of children with hypovitaminosis D were among those without medications for their cardiac defects’ complications. A plausible explanation might be that commencement of medications in those that need them will promote haemodynamic stability and thus make them less predisposed to malnutrition. There was no association between the previous number of hospital admissions and hypovitaminosis D. Studies done in children and adults without congenital heart disease reveal that frequent hospitalization and severity of illness were associated with hypovitaminosis D.^48,49^ Only about one-fourth of all the subjects had been on hospital admission in the last one year before recruitment and out of these, 81.5% had only been on admission once. No relationship was found in the present study and this may be attributed to the fact that majority (81.5%) of the subjects had only been on admission once. In developed countries, emergency room visits of children with congenital heart disease has reduced over time.^32^ This has been attributed to more refined medical therapy, higher resource devotion and medical attention to children with congenital heart defect. This however does not represent the reason for the lower hospital admission in the present study as facilities for surgical intervention in developing countries are not readily accessible and available. The lower rate of hospital admission in the present study might be attributed to poor health seeking behaviours, worsening economy

The median duration between the diagnosis of congenital heart disease among subjects and recruitment into the study was 25 months and almost one-fourth of subjects with congenital heart disease presented more than two years ago. Animasahun et al^50^ in Lagos State reported the mean age of children with cyanotic congenital heart disease who had not had surgery to be 47.4 months despite the fact that most cases of cyanotic congenital heart disease are diagnosed in the neonatal period. Majority of these children go for months without surgery. This shows the burden of congenital heart disease in Africa, especially West Africa where many have limited access to surgery due to the financial implications, inadequate health facilities and lack of expertise. This is in different from a report by Gilboa et al^10^ in the United States where mortality arising from congenital heart disease over a period seven years has reduced due to improved diagnostic capacities, surgical techniques and prenatal diagnosis.

Children with congenital heart disease had significantly lesser sunlight exposure compared with controls in the current study. Reduced physical activity in children with congenital heart disease has been attributed to physical fatigue, low self-efficacy and covert fears. Parents of children with congenital heart disease also over-restrict and impose unnecessary limitations there by reducing their physical activity due to under-estimation of their physical abilities, certain myths and beliefs about their condition and all these might result in inadequate sunlight exposure. After a careful literature search, there was no published report on sunlight exposure of children with congenital heart disease, hence the finding of the present study could not be compared. However, Stone^114^ and Voss^115^ compared the level of physical activity in children with congenital heart disease with healthy controls and no significant difference was found.

## Conclusions

The study shows prevalence of hypovitaminosis D is significantly higher in children with congenital heart disease when compared with that of apparently healthy controls while there is no difference in pattern between subjects with acyanotic and cyanotic CHD. There is a need for children with congenital heart disease to have periodic assessments of vitamin D level with need for vitamin D supplementation and dietary counselling on vitamin D rich foods among them

## Data Availability

All data produced in the present study are available upon reasonable request to the authors

## Acknowledgement

We thank participants and their caregivers for creating time to be part of the study. Our profound appreciates to doctors, nurses and Mr. Clement Akinsola (Statistician) who provide care and help during the analysis of the projects

